# Pan-oesophageal and spatially resolved genetic analysis reveals instability signature for earlier diagnosis of squamous cell cancer

**DOI:** 10.1101/2025.11.27.25340016

**Authors:** Aisha Yusuf, Shalini Malhotra, Maria O’Donovan, Ginny Devonshire, Sarah Killcoyne, Maryla H. Turkot, Andrzej Mroz, Malgorzata Lenarcik, Michal Mikula, Nastazja D. Pilonis, Jaroslaw Regula, Michal F. Kaminski, Karol Nowicki-Osuch, Wladyslaw Januszewicz, Rebecca C. Fitzgerald

## Abstract

Early detection of oesophageal squamous cell carcinoma (OSCC) significantly improves patients’ survival. Nonetheless, the availability of non-endoscopic, effective, and minimally invasive diagnostic approaches is limited. Here, we evaluated the utility of minimally invasive pan-oesophageal sampling using a capsule-sponge and aneuploidy detection using shallow whole genome sequencing (sWGS) for early detection of OSCC and precancerous intraepithelial neoplasia (IEN).

In the prospective arm, 200 participants underwent the capsule-sponge procedure, and we performed sWGS from 178 successfully collected specimens (89%). We combined newly developed genome-wide copy-number alteration (GW-CNA) score and copy number alterations (CNAs) at chromosomal arm level to measure global and local aneuploidy, respectively. Logistic regression model identified GW-CNA and CNAs on chromosomal arms 2q, 3q, 9p and 11q as key diagnostic predictors differentiating OSCC and IEN from healthy controls (AUC of 0.920 (95% CI: 0.907-0.933), accuracy: 0.888, sensitivity: 0.896, specificity: 0.887). The model outperformed histology-based diagnosis using H&E staining and p53 immunohistological assessment. Finally, the analysis of microdissected samples derived from retrospective endoscopic *en-bloc* resections, and spanning the entire pathological continuum of OSCC demonstrated stepwise increase in GW-CNA and CNAs of 2q, 3q, 9p and 11q, validating their biological significance.

This study demonstrates the high potential of combined pan-oesophageal sampling and sWGS aneuploidy analysis for early detection of OSCC and as a potential path to improved patients’ outcomes.

## INTRODUCTION

Despite substantial advances in cancer therapeutics, overall mortality rates for many gastrointestinal malignancies have scarcely improved over recent decades, mainly due to late-stage diagnosis. Oesophageal cancer exemplifies this challenge: most cases are detected at advanced stages, limiting curative options and resulting in a five-year survival rate of less than 20%^1,2^. Oesophageal squamous cell carcinoma (OSCC), which accounts for nearly 90% of oesophageal cancer cases globally, is especially prevalent in East Asia, Africa, South America, and Eastern Europe^3^. In 2020 alone, over 600,000 new OSCC cases were reported worldwide, a number that is projected to rise substantially in the coming years^4^.

OSCC arises *de novo* from the squamous epithelium through a stepwise carcinogenic pathway, progressing from low-grade intraepithelial neoplasia (LG-IEN) to high-grade intraepithelial neoplasia (HG-IEN), and ultimately to invasive carcinoma. Survival is strongly associated with stage and most patients present late^5,6^. By contrast, the diagnosis of IEN can achieve over 95% overall 5-year survival rate when combined with appropriate endoscopic therapy^7^. The stepwise progression to OSCC, the availability of endoscopic treatment at precancerous and early disease stages, and the survival benefits associated with early diagnosis offer an excellent opportunity for early detection and interception of this cancer at the pre-invasive and stage I phase of OSCC tumorigenesis.

Although endoscopy with biopsy remains the diagnostic gold standard for OSCC, its invasiveness, cost, and reliance on specialised infrastructure limit scalability for population-wide screening^8^. The limitations of endoscopic diagnosis are especially apparent in resource-poor setting of East Asia and Africa, where OSCC is endemic^9^. As a result, non-endoscopic approaches to OSCC diagnosis are of great interest. This can be exemplified by studies exploring the collection of saliva, exhaled breath condensates and liquid biopsies for the analysis of novel biomarkers, including gene methylation panels, microRNAs and multi-protein signatures^8^. Furthermore, machine learning-driven cytology has shown promise^10^. However, the clinical utility is often constrained by variable sensitivity, interpretive subjectivity, sampling bias, or limited mechanistic linkage of biomarkers to OSCC biology. The new generation of capsule sponge devices^11^ capable of broad oesophageal sampling offer an affordable, scalable, and unbiased sampling platform for molecular biomarker interrogation, particularly in light of the decreasing cost and growing availability of genome sequencing technologies.

Among candidate molecular biomarkers are copy number alterations (CNAs) - changes in the number of copies of genomic segments, that can range from focal events affecting single genes to large-scale chromosomal gains or losses. In OSCC, large-scale CNAs emerge early in squamous neoplasia tumorigenesis and contribute to tumour progression by altering gene dosage of key oncogenes and tumour suppressors^12–15^. Despite their biological relevance and potential for early detection, the efficacy of CNAs has not yet been investigated as an early diagnostic biomarker of OSCC either from endoscopic biopsies nor combined with minimally invasive, non-endoscopic screening strategies.

Here, we address this gap by applying shallow whole-genome sequencing (sWGS) to oesophageal cells collected via a minimally invasive capsule sponge from individuals at elevated risk of OSCC. To quantify global chromosomal instability, we developed a genome-wide copy number alteration (GW-CNA) burden metric, complemented by systematic profiling of recurrent chromosome-arm-level CNAs. Recognising that capsule sponge specimens represent a heterogeneous, pan-oesophageal cell population, we additionally analysed an independent cohort of microdissected endoscopic resection specimens to determine whether CNAs identified non-endoscopically reflect changes confined to dysplastic or malignant lesions, or whether they also arise from a wider field effect. These microdissected regions were precisely targeted and encompassed histologically defined stages of OSCC evolution, from normal epithelium through LG-IEN and HG-IEN to invasive carcinoma. This molecular validation framework enabled us to distinguish lesion-specific genomic alterations from broader field effects, anchoring these biomarkers within the natural history of OSCC and informing strategies for early detection and prevention.

## RESULTS

### Study design and patient cohort characteristics

The study was conducted in a Central European population, where OSCC remains the predominant histological subtype of oesophageal cancer. We used two complementary approaches. First, we recruited a prospective cohort of 200 individuals who underwent the capsule sponge procedure followed by endoscopy on the same day (Fig. 1A, table 1). Second, to understand the biological relevance and timing of the copy number aberrations identified from the sponge, we micro-dissected 110 specimens from a cohort of 32 patients, representing discrete stages of disease progression (Fig. 1B, Table 2).

**Figure 1:**
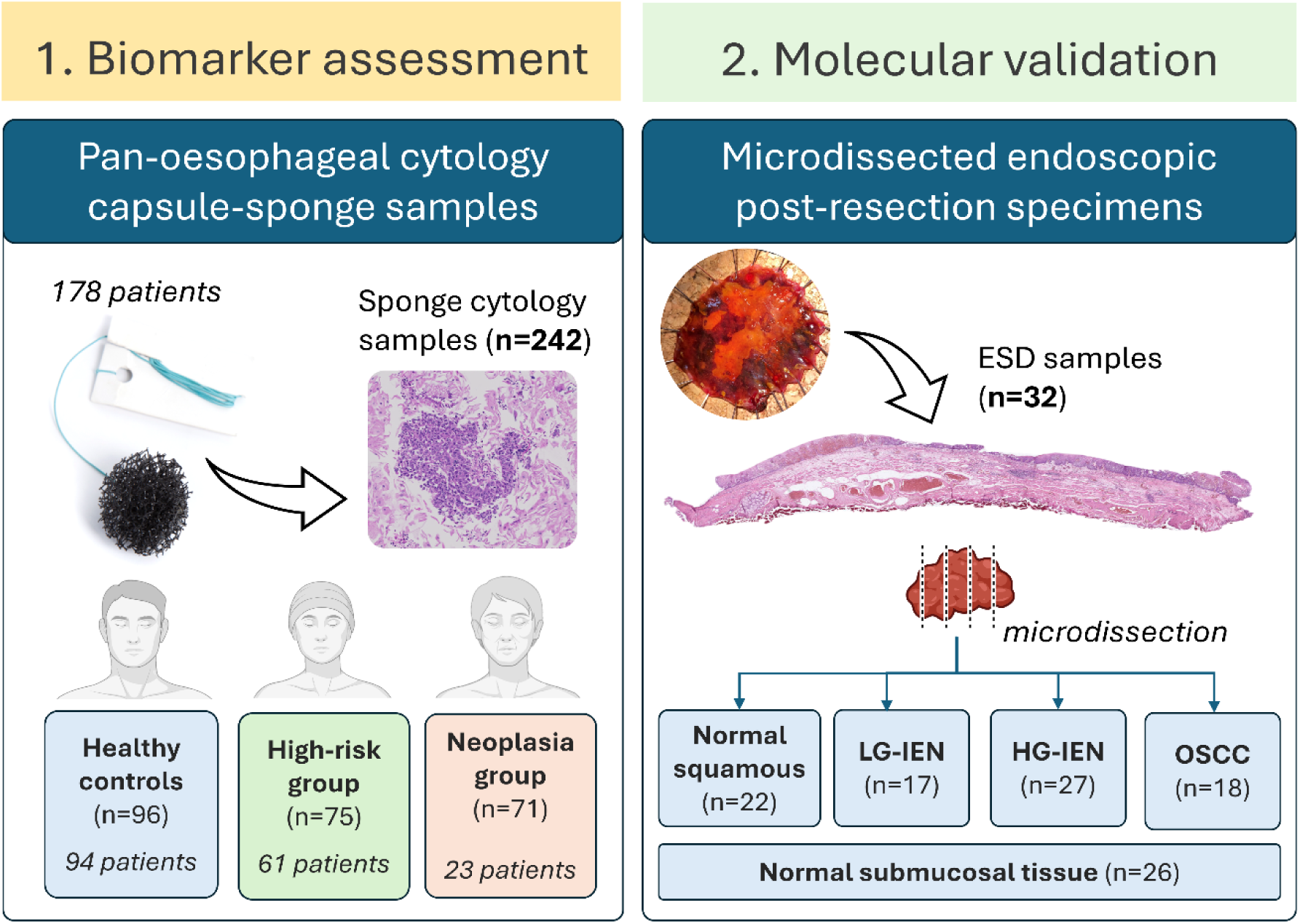
Study overview. Summary of prospective capsule sponge device cohort (1) and retrospective microdissection-based (2) components of the study. 1) Each group shows the number of participating individuals after exclusion. 2) Specimen we collected from 32 individuals and the number of samples used in the subsequent analysis is shown.

**Table 1:** Demographic and clinical characteristics of the prospective study cohort. Summary of demographic information, clinical data, and anthropometric measures for the final cohort of 178 participants: the healthy control group (n=94), high-risk individuals (n=61), and neoplasia group (patients with oesophageal IEN/early-cancer, n=23). The variables reported include mean age, sex distribution, body mass index (BMI), smoking status, lesion location along the oesophagus, and lesion type. Benign conditions represent all conditions identified through the histopathological assessment of endoscopic biopsies that are unrelated to the pro-gression of OSCC (e.g. oesophagitis, papilloma, acanthosis, necrosis, or koilocytosis). ^1^One participant initially enrolled in the high-risk group was subsequently reclassified to the neoplasia group following biopsy-confirmed proximal HG-IEN diagnosis.

| Characteristic | Healthy control group<br>(n = 94) | High-risk group<br>(n = 61) | Neoplasia group<br>(n = 23) |
| --- | --- | --- | --- |
| Age, years (mean ± SD) | 56.0 ± 13.6 | 64.4 ± 12.2 | 66.1 ± 9.3 |
| Male sex, n (%) | 51 (54.3%) | 42 (68.9%) | 11 (47.8%) |
| BMI, kg/m<sup>2</sup> (median [IQR]) | 26.2 [23.4–29.2] | 26.5 [24.7–28.9] | 23.9 [22.4–27.3] |
| Smoking (active/former), (%) | 49.5 | 80.5 | 70.6 |
| Lesion location, n (%) | N/A | Proximal: 1 (1.6%)<sup>1</sup> | Proximal: 11 (47.8%)<br>Middle: 7 (30.4%)<br>Distal: 5 (21.7%) |
| Histopathology, n (%) | Normal: 70 (74.5%)<br>Benign: 24 (25.5%) | Normal: 36 (59.0%)<br>Benign: 24 (39.3%)<br>HG-IEN: 1 (1.6%) | LG-IEN: 2 (8.7%)<br>HG-IEN/pTis: 8 (34.8%)<br>pT1a: 4 (17.4%)<br>pT1b: 7 (30.4%)<br>pT2: 2 (8.7%) |
| Number of scroll-sets collected per histological diagnosis | Normal: 72<br>Benign: 24 | Normal: 44<br>Benign: 29<br>HG-IEN: 2 | LG-IEN: 8<br>HG-IEN/pTis: 28<br>pT1a: 13<br>pT1b: 16<br>pT2: 6 |

**Table 2:** Demographic and clinical characteristics of the microdissected ESD molecular validation cohort. Thirty-two patients (mean age 62.0 ± 13.3 years; 62.5% male) underwent endoscopic submucosal dissection (ESD). Lesions were distributed across the oesophagus (proximal 34.4%, middle 40.6%, distal 25.0%). From these patients, 110 microdissected scroll-sets were profiled for genomic analyses (26 ST-NC, 22 ANSE, 17 LG-IEN, 27 HG-IEN, 18 OSCC).

|  |  |
| --- | --- |
| Age, years (mean ± SD; range) | 62.0 ± 13.3 (34–85) |
| Sex — Male, n (%) | 20 (62.5%) |
| Lesion location [Proximal, n (%)] | 11 (34.4%) |
| Lesion location [Middle, n (%)] | 13 (40.6%) |
| Lesion location [Distal, n (%)] | 8 (25.0%) |
| Histopathology (index lesion) [LGD, n (%)] | 4 (12.5%) |
| Histopathology (index lesion) [HGD, n (%)] | 9 (28.1%) |
| Histopathology (index lesion) [OSCC, n (%)] | 19 (59.4%) |

**Table 2: Demographic and clinical characteristics of the microdissected ESD molecular validation cohort.**
|  |  |
| --- | --- |
| Submucosal tissue [negative control (ST-NC)] | 26 (23.6%) |
| Adjacent normal squamous epithelium (ANSE) | 22 (20.0%) |
| Low-grade intraepithelial neoplasia (LG-IEN) | 17 (15.45%) |
| High-grade intraepithelial neoplasia (HG-IEN) | 27 (24.55%) |
| Invasive OSCC | 18 (16.36%) |

In the prospective cohort, capsule sponge samples collected from 178 individuals were analysed. 14 (7%) initially recruited individuals were unable to swallow the device, and 8 (4.6% out of 186) capsule sponge samples were not analysed due to technical failures (Suppl. Fig 1). Depending on their clinical characteristics prior to capsule sponge administration, patients were assigned to three experimental groups: healthy controls (94), high-risk group (61) and neoplasia group (23). The neoplasia group comprised patients with known early oesophageal squamous neoplasia undergoing evaluation for endoscopic resection (Table 1), while the high-risk group consisted predominantly of individuals under endoscopic surveillance following definitive treatment for head and neck cancers (oral cavity, hypopharyngeal, pharyngeal, or laryngeal carcinoma) who are known to be at increased with for oesophageal cancer, alongside a smaller subset with prior definitive endoscopic therapy for early OSCC. To offset the relative scarcity of neoplastic cases, we implemented a pre-specified stratified design that allocated multiple scroll-sets per neoplasia specimen (typically three), with proportionally fewer scroll-sets prepared from the high-risk and control groups, thereby maximising analytical depth while preserving group representation (Fig. 1, Suppl. Fig. 1). A total of 242 scroll-sets were sequenced; 96 from healthy controls, 75 from high-risk participants, and 71 from the neoplasia group (Table 1). Participants swallowed the capsule sponge device on the same day as their scheduled endoscopy. Diagnostic biopsies from the endoscopy procedure, or final staging of the endoscopically resected specimens, served as the gold-standard reference.

Participants in the high-risk and neoplasia groups were significantly older than controls (mean age: 64.4 ± 12.2 and 66.1 ± 9.3 years vs 56.0 ± 13.6 years; p < 0.001) and more likely to be current or former smokers (80.5% and 70.6% vs 49.5%; p = 0.002) (Table 1). Median BMI was 26.2 kg/m² in controls, 26.5 kg/m² in the high-risk group, and 23.9 kg/m² in the neoplasia group; the proportion of male participants was highest in the high-risk group (68.9%) (Table 1).

Endoscopic biopsy confirmed normal oesophageal histology in 106 of 178 participants (59.6%), benign inflammatory changes such as oesophagitis, papilloma, or acanthosis in 48 (27.0%), and IEN or early squamous carcinoma in 24 (13.5%) (Table 1). This includes one participant initially enrolled in the high-risk group who was reassigned to the neoplasia group after histological confirmation of proximal HG-IEN.

We established an independent retrospective molecular validation cohort of 32 patients who underwent endoscopic submucosal dissection, from which 110 microdissected samples were profiled (Table 2). The cohort had a mean age of 62.0 ± 13.3 years (range 34-85) and was 62.5% male. Lesions were distributed across the proximal (34.4%), middle (40.6%), and distal (25.0%) oesophagus. Histopathological assessment of the index lesions identified 4 LGD (12.5%), 9 HGD (28.1%), and 19 OSCC (59.4%). The microdissected material comprised 26 submucosal tissue negative controls (ST-NC; included as true tissue controls), 22 adjacent normal squamous epithelia (ANSE; included to assess potential field effects), 17 LG-IEN, 27 HG-IEN, and 18 OSCC specimens.

### Genome-wide chromosomal instability emerges at the earliest stages of squamous neoplasia

Next, we performed sWGS of 242 scroll sets obtained using capsule sponge and stratified scroll-sets using histopathological diagnosis after endoscopy as reference. When split by the experimental group, we observed a near-diploid genomic landscape in histologically normal (n = 116) or benign inflammatory epithelium (n = 53) (Table 1, Fig. 2). These samples were characterised by tightly clustered log_₂_ copy-number values across autosomal chromosome arms (median arm-level s.d. ≈ 0.02; Fig. 2). By contrast, scroll-sets collected from patients diagnosed with squamous neoplasia (LG-IEN, HG-IEN, OSCC) at endoscopy (n = 73) exhibited significantly greater chromosomal dispersion (median s.d. = 0.032), indicative of widespread genomic instability (Kruskal-Wallis FDR-adjusted p = 1.5 × 10⁻⁶). Post-hoc Dunn tests confirmed increased dispersion relative to benign (adjusted p < 1 × 10⁻⁴) and normal (adjusted p < 1 × 10⁻⁶) epithelium.

**Figure 2:**
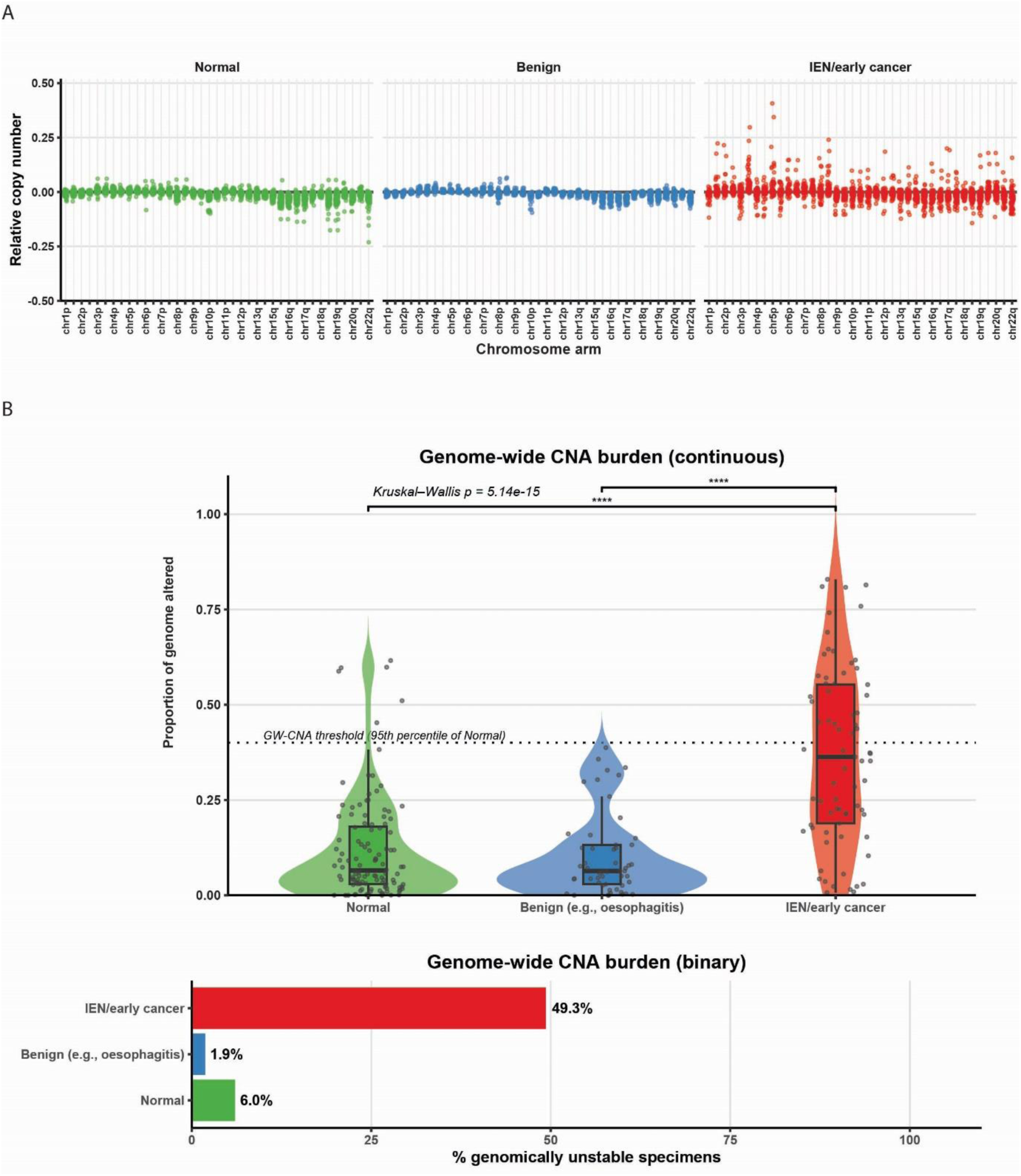
Chromosomal copy-number variability distinguishes normal, benign, and neoplastic epithelium. A) Scatter plots of log₂-transformed relative copy number (RCN) values across autosomal chromosome arms (1p–22q) are shown for individual scroll sets from capsule sponge specimens, stratified by histological diagnosis after endoscopy: normal epithelium (n = 116, green), benign epithelium (e.g., oesophagitis; n = 53, blue), and intraepithelial neoplasia/early cancer (IEN; n = 73, red). Each point corresponds to a sample’s RCN for a given chromosome arm. B) Top, distribution of GW-CNA burden, defined as the proportion of the genome encompassed by significant chromosome-arm gains or losses, across Normal (green, n = 116), Benign (blue, n = 53; e.g., oesophagitis), and IEN/early cancer (red, n = 73) groups. The dotted line indicates the genomic instability threshold (95th percentile of Normal). Kruskal–Wallis p value is shown; significant pairwise differences versus IEN/early cancer are annotated (FDR-adjusted Dunn’s tests). Bottom, proportion of specimens exceeding the threshold and therefore classified as genomically unstable.

To quantify this instability, we computed the genome-wide copy-number alteration (GW-CNA) burden, defined as the fraction of the genome encompassed by significant chromosome-arm gains or losses. GW-CNA burden rose markedly across the histological continuum (median [IQR]: normal, 0.07 [0.15]; benign, 0.06 [0.10]; neoplasia, 0.36 [0.36]; Fig. 2B), with a with a global Kruskal-Wallis FDR-adjusted p = 5.0 × 10⁻¹³ and significant pairwise differences (neoplasia diagnosis versus benign and normal both adjusted p-value < 1 × 10⁻¹¹). The substantial increase highlights the early acquisition and accumulation of chromosomal imbalance during oesophageal carcinogenesis.

For clarity and practical applicability, we established a binary genomic instability threshold at the 95th percentile of GW-CNA in normal controls (threshold = 0.365). Using this definition, 49.3% of dysplasia/early-cancer specimens were classified as genomically unstable compared to only 6.0% of normal and 1.9% of benign samples (χ² = 68.5, P < 0.001; Fig. 2B). Collectively, these data establish GW-CNA (and its binary derivative) as robust, objective measures of genome-wide chromosomal instability emerging at the earliest stages of oesophageal squamous neoplastic progression.

To identify key, chromosome specific, genomic predictors of dysplasia and early cancer, we applied elastic net regularisation within a generalised linear model framework, analysing relative copy number values across 39 chromosomal arms to be used alongside the GW-CNA burden. By focusing on features with consistent relative risk estimate (see methods), we identified five key predictors: chromosomal arms 2q, 3q, 9p and 11q, and the GW-CNA burden metric (Fig. 3A and Suppl. Fig. 2).

**Figure 3:**
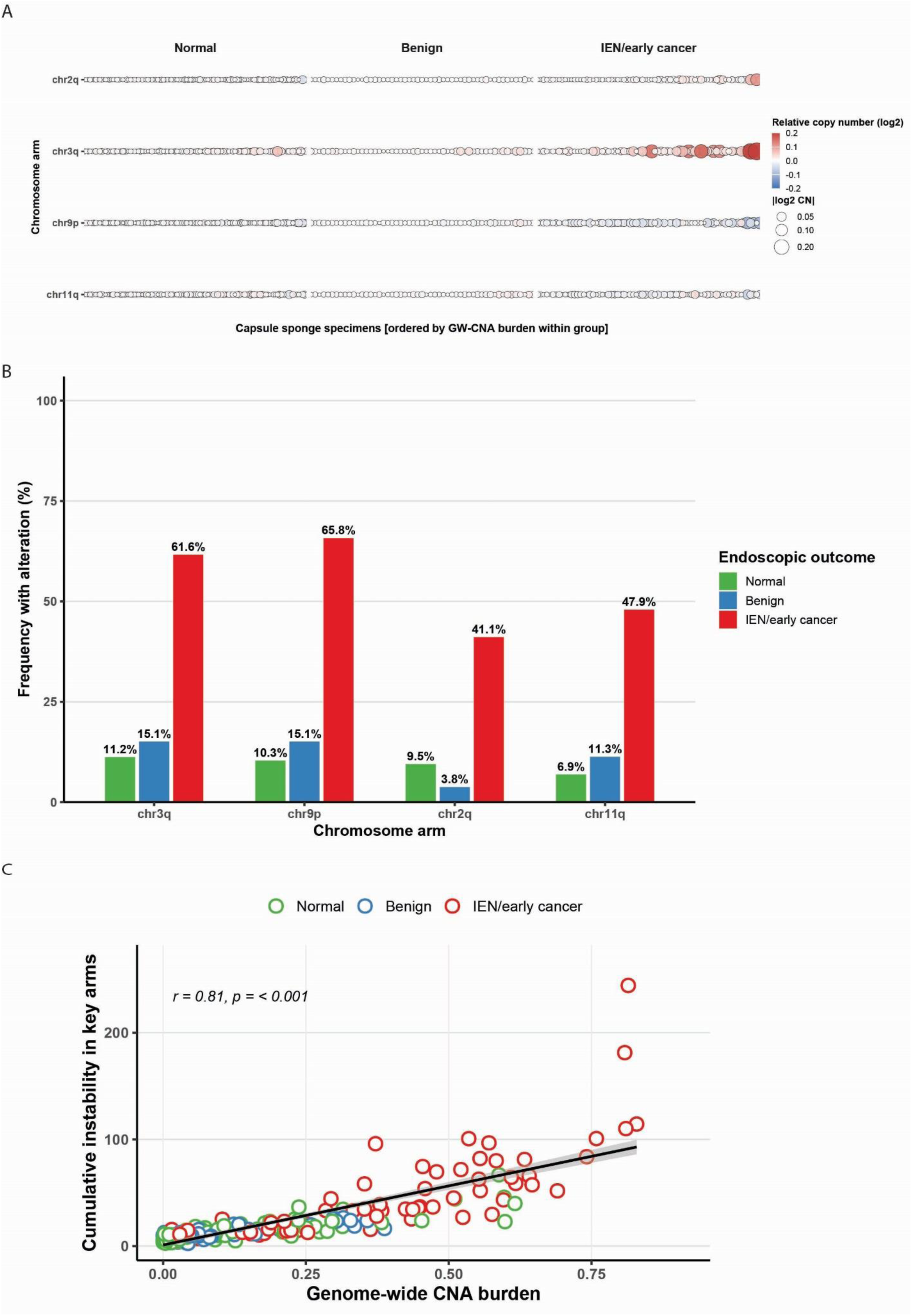
Arm-level copy number alterations in capsule sponge scroll-sets. A) Log₂-transformed relative copy number values across four key chromosomal arms (2q, 3q, 9p, 11q) in capsule sponge scroll-sets, stratified by endoscopic outcome: normal epithelium (green, n = 116), benign epithelium (blue, n = 53; including oesophagitis, papilloma, acanthosis, necrosis, or koilocytosis), and intraepithelial neoplasia/early cancer (IEN; red, n = 73). Within each outcome group, scroll-sets are ordered left-to-right by genome-wide CNA burden. Each point represents a chromosomal arm within a scroll-set, with size proportional to the magnitude of the copy number change (|log₂|) and colour indicating direction and extent (red, gain; blue, loss; white, near-diploid). B) Proportion of scroll-sets with copy number alterations (gain or loss) in each diagnostic group across the same four chromosomal arms, expressed as percentages. C) Scatter plot demonstrating the strong positive correlation (Pearson’s r = 0.814, p < 0.001) between genome-wide copy number alteration (GW-CNA) burden and cumulative instability in key chromosomal arms (2q, 3q, 9p, and 11q) from Capsule Sponge specimens.

Specifically, arm 3q alterations occurred in 61.6% (45/73) of neoplasia scrolls-set versus 15.1% (8/53) in benign and 11.2% (13/116) in normal scroll-sets (χ² = 62.5, P = 2.6 × 10⁻¹⁴) (Fig. 3B). Similarly marked differences were observed for 9p (65.8%, 48/73 vs. 15.1%, 8/53 and 10.3%, 12/116; χ² = 73.8, P = 9.6 × 10⁻¹⁷), 2q (41.1%, 30/73 vs. 3.8%, 2/53 and 9.5%, 11/116; χ² = 39.7, P = 2.3 × 10⁻⁹), and 11q (47.9%, 35/73 vs. 11.3%, 6/53 and 6.9%, 8/116; χ² = 50.1, P = 1.3 × 10⁻¹¹). At least one alteration across these key chromosomal arms was observed in the majority (86.3%, 63/73) of neoplastic scroll-sets, compared to 32.1% (17/53) of benign and 25.0% (29/116) of normal scroll-sets, further emphasizing their collective importance (Fig. 3B). There was also a strong positive correlation between cumulative alterations in these key chromosomal arms and GW-CNA burden (Pearson’s r = 0.81, P < 0.001; Fig. 3B), suggesting an association between localised chromosomal instability and broader genome-wide disruption.

### Logistic Regression Model to detect dysplasia and cancer from pan-oesophageal samples

To evaluate the diagnostic potential of genome-wide chromosomal instability metrics combined with specific chromosomal alterations in a non-endoscopic pan-oesophageal sample, we developed logistic regression models incorporating both continuous and binary measures of GW-CNA burden, alongside key chromosomal arms (2q, 3q, 9p, and 11q). Focusing on individual scroll-sets, in the comparison of neoplasia diagnosis against benign or normal endoscopic outcomes, the continuous GW-CNA burden model achieved an AUC of 0.920 (95% CI: 0.907-0.933), with an accuracy of 0.888, sensitivity of 0.896, and specificity of 0.887 (Figure 4, Suppl. Table 1). The binary burden model demonstrated comparable performance (AUC: 0.922, 95% CI: 0.908-0.935), with a slightly higher sensitivity (0.899) and specificity (0.904).

**Figure 4.**
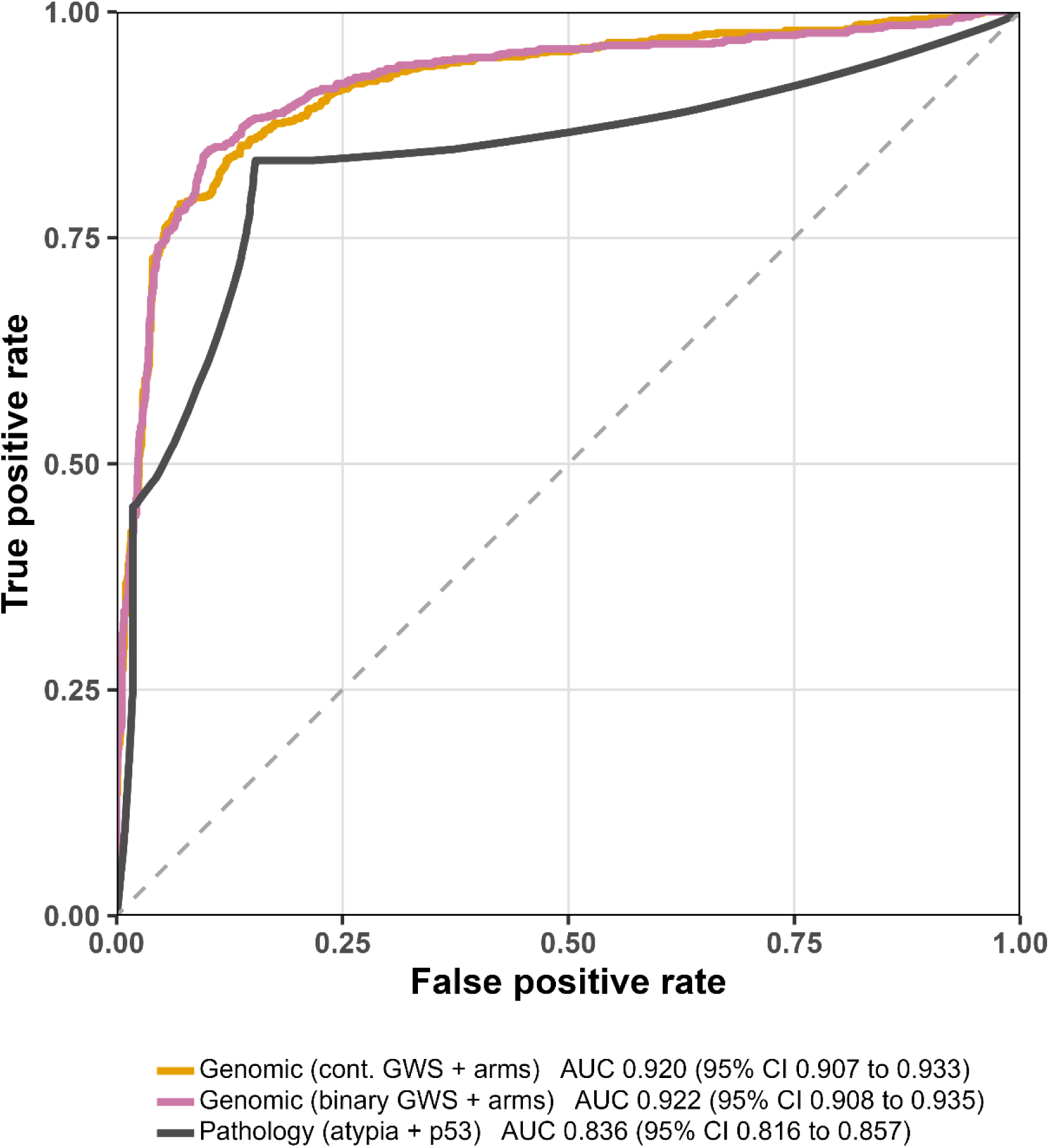
Diagnostic performance of genomic and pathology-based models. Receiver operating characteristic (ROC) curves comparing the ability of genomic models and pathology-based markers to distinguish between intraepithelial neoplasia (IEN) or early oesophageal squamous cancer and non-neoplastic outcomes. The genomic models incorporate either continuous (orange line) or binary (magenta line) genome-wide copy number alteration (GW-CNA) burden, alongside chromosomal alterations on 2q, 3q, 9p, and 11q. The pathology-based model combines capsule sponge-derived squamous atypia and p53 immunohistochemistry (black line).

To benchmark the genomic models, we compared them to a pathology-based diagnostic approach, in which capsule sponge samples were evaluated for squamous atypia by two expert histopathologists and for p53 status by immunohistochemistry (IHC) (Methods). This combination achieved an AUC of 0.836 (95% CI: 0.816-0.857), with a sensitivity of 0.851 and specificity of 0.831 (Figure 4, Suppl. Table 1). Despite a high positive predictive value (PPV 0.923), this strategy missed nearly 15% of IEN and early cancer cases and misclassified approximately 17% of benign samples, likely reflecting reactive epithelial changes. In contrast, the genomic model incorporating continuous GW-CNA burden reduced these error rates to ∼10% and ∼11%, respectively, while achieving an AUC of >0.9.

Analysis of the logistic regression coefficients (Suppl. Tables 2A and 2B) identified gains in 3q as a strong predictor of increased risk for IEN and early cancer (continuous model: β = 81.8, p = 0.0006; binary model: β = 87.0, p = 0.0002). Losses in 9p were also consistently associated with elevated risk (continuous: β = −140.0, p < 0.0001; binary: β = −147.0, p < 0.0001). These findings align with previously implicated regions of chromosomal instability in oesophageal squamous neoplasia, though the current analysis does not resolve gene-level alterations.

Unlike the capsule sponge, which captures a pan-oesophageal sample and precludes precise pathological attribution, micro-dissected endoscopic submucosal dissection (ESD) specimens provide spatial resolution to determine the extent to which copy number changes coincide with disease stage and to confirm any field-effects (Fig. 5A). We focused on the key copy-number alteration (CNA)-related biomarker candidates identified in the capsule sponge model (2q, 3q, 9p, 11q and genome-wide CNA burden) and analysed them across five pathological entities: submucosal tissue negative control (ST-NC), adjacent normal squamous epithelium (ANSE), LG-IEN, HG-IEN and OSCC (Fig. 5A).

**Figure 5:**
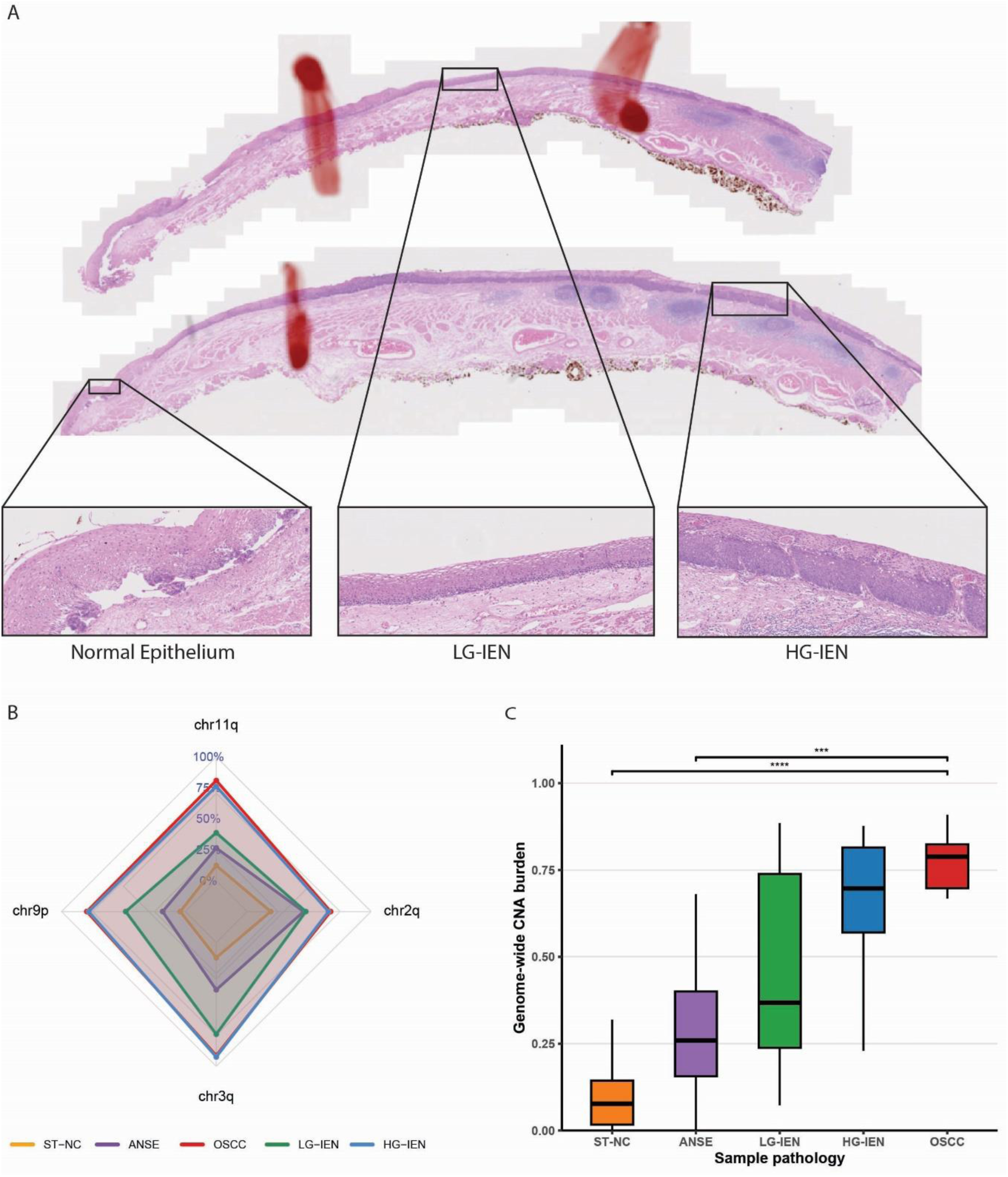
Genetic analysis of microdissected endoscopic submucosal dissection (ESD) specimen. A) Representative haematoxylin and eosin (H&E)–stained section of a formalin-fixed paraffin-embedded (FFPE) oesophageal endoscopic submucosal dissection (ESD) specimen from a patient with regions of low- (LG-IEN) and high-grade intraepithelial neoplasia (HG-IEN). Regions selected for microdissection are highlighted in the insets. B) Radar plots of the proportion of microdissected ESD specimens with copy number alterations in four key chromosomal arms (2q, 3q, 9p, 11q). Each polygon represents the mean proportion of altered segments within a pathology group: submucosal tissue negative control (ST-NC, orange), adjacent normal squamous epithelium (ANSE, purple), LG-IEN (green), HG-IEN (blue) and OSCC (red). C) Boxplot of GW-CNA burden, defined as the as the proportion of the genome encompassed by significant chromosome-arm gains or losses across microdissected endoscopic submucosal dissection specimens: ST-NC (n = 26, orange), adjacent normal squamous epithelium (ANSE, n = 22, purple), low-grade dysplasia (LGD, n = 17, green), high-grade dysplasia (HGD, n = 27, blue), and OSCC (n = 18, red). GW-CNA burden increased progressively from precancerous to malignant stages. Statistical significance was assessed by Kruskal–Wallis test (p < 0.001), with selected post-hoc comparisons adjusted by Hochberg correction (*p < 0.05, **p < 0.01, ***p < 0.001).

2q gains showed a gradual increase from 15.4% (4/26) in ST-NC to 40.9% (9/22) in ANSE and 47.1% (8/17) in LG-IEN, further rising to 55.6% (15/27) in HG-IEN and 61.1% (11/18) in OSCC, suggesting that 2q alterations accumulate progressively rather than marking a discrete transition. 3q gains emerged as the earliest and most prominent alterations, detected in 7.7% (2/26) of ST-NC and increasing to 31.8% (7/22) in ANSE, before rising sharply to 70.6% (12/17) in LG-IEN (p < 0.01, adjusted) and 88.9% in both HG-IEN (24/27) and OSCC (16/18) (Figure 5B). Hence, 3q gains are significantly more frequent in HG-IEN than in ANSE (p < 0.01, adjusted). 9p losses followed a progressive trajectory and became particularly pronounced in mid-stage disease. They were detected in 3.8% (1/26) of ST-NC, rising to 18.2% (4/22) in ANSE and 47.1% (8/17) in LG-IEN, with a marked increase to 74.1% (20/27) observed in HG-IEN (p < 0.01, adjusted vs. ST-NC); their frequency then remained high in OSCC (72.2%; 13/18). In contrast, 11q gains were more prominent at later stages, escalating from 11.5% (3/26) in ST-NC to 22.7% (5/22) in ANSE and 41.2% (7/17) in LG-IEN, before surging to 74.1% (20/27) in HG-IN (p < 0.05, adjusted vs. ANSE) and remaining high at 83.3% (15/18) in OSCC (Figure 5B). Across all stages, at least one alteration in these four arms (2q, 3q, 9p, 11q) was detected in 19.2% of ST-NC, 63.6% of ANSE, 88.2% of LG-IEN, 96.3% of HG-IEN, and 88.9% of OSCC.

When considering the GW-CNA burden as a holistic measure of chromosomal instability in the ESD samples, there was a progressive increase across pathology stages (ST-NC < ANSE < LG-IEN < HG-IEN < OSCC), consistent with the changes seen in capsule sponge samples (Figure 5). Median values increased from 0.077 (interquartile range [IQR]: 0.017-0.144) in ST-NC to 0.652 (IQR: 0.507-0.710) in OSCC, with significant pairwise increases between ST-NC and LG-IEN (p = 0.005), LG-IEN and HG-IEN (p = 0.048), HG-IN and OSCC (p = 0.023), and LG-IN and OSCC (p = 0.002, all adjusted) (Figure 5C).

## DISCUSSION

In this paper, we present an integrated genomic model for OSCC early detection based on a genome wide and chromosomal arm-level measure of genomic instability that has biological validity since the changes are specific to areas of phenotypic dysplasia and show increasing signal with the severity of disease.

The progressive increase in the GW-CNA burden from normal and benign tissues to dysplasia and early OSCC that aligns with established models of escalating chromosomal instability in squamous neoplasia suppressors^12–15^. Furthermore, the strong correlation between cumulative CNA alterations in 3q, 9p, 2q, and 11q and the GW-CNA burden further underscores the interconnected roles of focal and genome-wide genomic instability in OSCC progression.

In non-endoscopic capsule sponge samples, our combined CNA signature achieved high diagnostic performance (AUC up to 0.922, with approximately 90% sensitivity and specificity), surpassing conventional cytology-based assessments, namely squamous atypia and p53 IHC, which misclassified 16% of dysplastic lesions and 15% of non-dysplastic cases in our cohort. These findings highlight the inherent limitations of morphology-based diagnostics, which are prone to observer variability and false positives arising from reactive atypia, particularly in patients with prior radiotherapy or inflammatory conditions. This is in contrast to squamous atypia plus p53 IHC which demonstrated near-perfect accuracy in a proof-of-concept study in high-incidence Iranian populations who had not had any prior radiotherapy^16^, our evaluation in a more heterogeneous European cohort underscores the critical need for objective molecular biomarkers that provide consistency across diverse clinical settings.

Alternative molecular approaches for non-endoscopic OSCC screening have shown promise but exhibit significant limitations. Gene methylation panels achieve high specificity (77-99%) but variable sensitivity (9-50%), limiting their clinical utility^17–19^. Protein-based assays, while promising, particularly those incorporating multi-marker signatures, face challenges in detecting early dysplasia as well as distinguishing dysplastic from benign tissue due to overlapping protein expression and subjective interpretation of staining^20,21^. Recent advances in machine learning-driven cytology have yielded strong classification metrics (AUC = 0.927 in community-based screening; AUC = 0.951 in nationwide studies)^10,22^. However, while these models leverage large-scale computational filtering, the biological significance of many algorithm-selected features remains uncertain, complicating their integration into mechanistically guided frameworks. In contrast, our CNA-based model achieved comparable diagnostic performance while directly measuring chromosomal instability, an established hallmark of carcinogenesis, offering a more biologically grounded alternative for OSCC detection.

Our analysis demonstrated frequent CNA alterations in chromosomal arms previously associated with OSCC diagnosis (3q, 9p, 2q, and 11q). For example, we observed that gains in 3q emerged as the earliest and most frequent alteration and were present across all stages of disease, consistent with prior studies showing that 3q amplifications, which include oncogenes such as SOX2 and PIK3CA, play an early and pivotal role in OSCC tumorigenesis^12,13,23^. SOX2 amplification has been correlated with epithelial stemness and proliferative signalling, whereas PIK3CA alterations have been linked to lymph node metastasis and advanced disease^24,25^. Other predictive chromosomal arms harbour well-known cancer genes, including CDKN2A (chromosome 9p), CCND1, CTTN, and FGF3/4/19 (chromosome 11q), and NFE2L2 (chromosome 2q).

This study has several strengths, including its focus on an underrepresented European OSCC population and the integration of spatially resolved microdissected specimens to verify the biological relevance of genomic alterations. However, some limitations warrant consideration. The modest sample size of dysplastic cases underscores the need for a larger prospective validation study. While sWGS provides a cost-effective and scalable approach for CNA detection, higher-resolution sequencing could further delineate driver genes within altered regions in the exfoliative oesophageal specimens. Finally, real-world implementation will require standardised sample processing and data analysis protocols to minimise technical variability, particularly in multi-center settings.

Overall, the promising performance of our genomic classifier (AUC up to 0.928), suggests that this approach could serve as a triage tool to identify high-risk individuals for confirmatory endoscopy, optimising healthcare resources in populations with lower OSCC prevalence. Although our findings are encouraging further prospective, large-scale studies are needed to validate these biomarkers across diverse settings. The ongoing ANGELA trial (NCT06418516) and other prospective studies will be critical in assessing real-world performance and generalisability.

## METHODS

### Cohort description and sample collection

Samples were obtained from two independent investigations: the Early Detection and Surveillance of Oesophageal Cancer with Cytosponge and Nucleic Acid Biomarkers (EDEN) study, conducted at the Maria Sklodowska-Curie National Research Institute of Oncology, and the Oesophageal Squamous Cell CARcinoma (OSCAR) study, conducted at Cambridge University Hospitals NHS Foundation Trust. Both studies received institutional ethics approval (OSCAR Research Ethics Committee No. 15/EE/0226; EDEN Ethics Committee No. 09/PB/2019). The EDEN study is registered at ClinicalTrials.gov (NCT04192695). International transfer of biospecimens from EDEN was performed under a formal collaboration agreement.

Participants were prospectively assigned to one of three predefined clinical categories:

i. **Healthy controls**: individuals without known oesophageal disease;
ii. **High-risk group**: individuals under endoscopic surveillance due to established risk factors for OSCC, including a prior history of head and neck squamous cell carcinoma or other predisposing conditions;
iii. **Neoplasia group**: individuals with histologically confirmed intraepithelial neoplasia (IEN; low- or high-grade) or early OSCC.

### Endoscopy and biopsy acquisition

All endoscopic examinations were performed by consultant gastroenterologists as part of clinical care, typically on the same day as Capsule sponge collection or within 7 days. High-definition video endoscopes (Olympus GIF-H180, GIF-H190, or Evis X1) were used under topical anaesthesia (lignocaine spray), intravenous conscious sedation (midazolam ± fentanyl), or general anaesthesia, as appropriate.

The oesophagus was examined using high-definition white light imaging, with adjunct narrow-band imaging and/or 2.0% Lugol’s iodine chromoendoscopy. Macroscopically abnormal areas were biopsied. Lesions <20 mm were sampled with two biopsies (diagnostic and research); larger lesions were sampled with three to four biopsies. In the absence of visible lesions, two biopsies of normal-appearing mid-oesophageal mucosa were obtained for research in high-risk participants. Specimens were snap-frozen or formalin-fixed and paraffin-embedded. Where indicated, patients underwent endoscopic mucosal resection (EMR), endoscopic submucosal dissection (ESD), or surgery; resection specimens served as the reference standard.

### Capsule sponge procedure

Participants fasted for ≥4 h. A tethered, gelatin-coated Cytosponge (Medtronic, Dublin, Ireland) was swallowed under direct supervision. After ∼5 min to allow sponge expansion in the oesophagus, the device was withdrawn by traction on the tether to recover exfoliated oesophageal cells. The sponge head was immediately placed in BD SurePath preservative medium (BD Diagnostics, Franklin Lakes, NJ, USA) and transported to the central laboratory. Adverse events and deployment failures were recorded.

### Capsule sponge processing and FFPE clot generation

Samples were processed at the Addenbrooke’s Hospital Tissue Bank (Cambridge, UK) using a standard protocol^26–29^. Upon receipt, samples were stored at 4 °C and processed within 3 days (up to 7 days).

Cellular material was released by mechanical dissociation, pelleted by centrifugation, and converted into FFPE “cell clots” using a plasma-thrombin method. Each FFPE block was sectioned into predefined “scroll-sets”, each consisting of: (i) an H&E-stained slide; (ii) an adjacent p53 immunohistochemistry (IHC) slide; and (iii) four 10 µm scrolls for DNA extraction and shallow whole-genome sequencing (sWGS). This ensured correlation between morphology, p53 status and genomic profiling. When necessary, multiple scroll-sets were prepared from the same FFPE block to address specific analytical requirements or enrich sample representation.

### Cytopathology and p53 immunohistochemistry

H&E-stained slides were reviewed by two expert histopathologists blinded to endoscopic findings (SM and MoD). Cytology was assigned using a classification analogous to the Bethesda System: normal; benign/reactive changes; atypical squamous cells (ASC; strongly suggestive of IEN); or atypical squamous cells of undetermined significance (ASCUS), indicating abnormalities insufficient for definitive IEN. Discrepant calls were resolved by consensus review.

Adjacent sections underwent p53 IHC on a Leica Bond-Max automated stainer (Leica Biosystems, Milton Keynes, UK) using the Bond Polymer Refine Detection Kit (cat. no. DS9800) and anti-p53 monoclonal antibody DO7 (Leica Biosystems, cat. no. NCL-L-p53-D07; 1:50 dilution). HET1A served as a positive control for each run. Slides with one or more squamous cells showing nuclear staining intensity of p53 comparable to that of the positive control slides were considered p53-positive. In clinical practice, equivocal p53 IHC patterns (e.g., patchy, or weak nuclear staining) are often conservatively reported as positive to minimise the risk of false negatives. Slides without nuclear staining were scored as p53-negative.

### Sampling strategy and sequencing

To increase analytical depth in clinically informative strata, a tiered sampling design was used. For participants in the neoplasia group, an average of three independent scroll sets were generated from each Capsule sponge FFPE block. A randomly selected subset of high-risk participants contributed two scroll-sets. Remaining high-risk participants and all healthy controls contributed one scroll-set. Each scroll-set was processed independently through DNA extraction, library preparation, and sequencing.

### DNA extraction

DNA was extracted from four 10 µm FFPE scrolls per library using the GeneRead DNA FFPE Tissue Kit (Qiagen, Hilden, Germany; cat. no. 180134), according to the manufacturer’s protocol. Sections were deparaffinised and digested with Proteinase K at 56 °C overnight. DNA was eluted twice in 15 µL Buffer ATE. DNA concentration was measured using a Qubit 2.0 fluorometer with the dsDNA High Sensitivity Assay Kit (Thermo Fisher Scientific, cat. no. Q32851). DNA integrity was assessed on an Agilent 4200 TapeStation (Agilent Technologies, Santa Clara, CA, USA) using Genomic DNA ScreenTape (cat. no. 5067-5365). Libraries were prepared from samples with DIN ≥3.

### Library preparation, pooling, and shallow whole-genome sequencing

Libraries were prepared using the NEBNext Ultra II FS DNA Library Prep Kit for Illumina (New England Biolabs, Ipswich, MA, USA; cat. no. E7805). Genomic DNA was fragmented at 37 °C for 5 min to generate ∼300-350 bp inserts, followed by end repair, 5′ phosphorylation, and dA-tailing. NEBNext Adaptors for Illumina were ligated (1:10 dilution for <50 ng input). Adaptor-ligated DNA was purified using 0.8× AMPure XP beads (Beckman Coulter, cat. no. A63880).

Libraries were amplified with NEBNext Multiplex Oligos for Illumina (Dual Index Primers Set; NEB, cat. no. E6440S). Seven PCR cycles were used for ≥50 ng input and nine cycles for ≤50 ng input. A 0.9× AMPure XP clean-up removed primer dimers. Fragment size (∼350-500 bp) and adaptor dimer absence were confirmed on an Agilent 4200 TapeStation using High Sensitivity D1000 ScreenTape (cat. no. 5067-5584). Libraries were quantified using the Qubit dsDNA Broad Range Assay (Thermo Fisher Scientific, cat. no. Q32853), normalised to target ∼1-2× genomic coverage per library, and pooled.

Balanced representation across pooled libraries was confirmed on an Illumina MiSeq Nano run (Illumina, San Diego, CA, USA). Final sequencing was performed on an Illumina NovaSeq 6000, generating 150 bp paired-end reads. Read counts per library were re-assessed post hoc to confirm uniform shallow coverage.

### Archival tissue cohort and laser-free microdissection

To enable spatially resolved comparison with Capsule sponge data, archival FFPE ESD tissue blocks were obtained from 32 patients (25 EDEN, 7 OSCAR). Blocks spanned adjacent normal squamous epithelium (ANSE), low-grade IEN (LG-IEN), high-grade IEN (HG-IEN), early OSCC, and uninvolved submucosal tissue. Submucosal tissue with intact submucosa uninvolved by tumour (ST-NC) was used as a diploid reference.

In total, 130 FFPE blocks were reviewed. Each block was sectioned into 4 µm slices for haematoxylin and eosin (H&E) staining and sequential 10 µm scrolls unstained sections (10 to 25) for DNA extraction. Two consultant gastrointestinal pathologists confirmed diagnosis, delineated regions of interest, and estimated cellularity of lesions. Only blocks with ≥20% cellularity in regions of interest were retained to minimise stromal contamination and ensure sufficient DNA yield.

Prior to microdissection, unstained slides were deparaffinised in xylene (∼5 min), rehydrated through two 3-min immersions in 99% ethanol, and air-dried completely. Regions of interest, including adjacent normal squamous epithelium (ANSE), low-grade IEN (LG-IEN), high-grade IEN (HG-IEN) and OSCC, were delineated on matched H&E slides. Submucosal tissue (ST-NC) was also marked in selected blocks. Guided by these annotations, manual microdissection was performed under a stereomicroscope (Leica MZ6, Leica Microsystems, Wetzlar, Germany). Target regions were excised using a sterile 21 G needle into microcentrifuge tubes containing 70% ethanol, vortexed, pelleted at 1,000 × g for 1 min, ethanol removed, and samples air-dried. DNA extraction, library preparation, and sequencing of microdissected tissue were performed as described for Capsule sponge specimens.

### Bioinformatic processing

Raw sequencing reads from Capsule sponge and microdissected tissue libraries underwent the same computational pipeline. Quality control was performed with FastQC (bioinformatics.babraham.ac.uk\projects\fastqc). Reads were aligned to the GRCh37 human reference genome using BWA-MEM v0.7.17 (Li and Durbin, 2009). PCR duplicates were marked using Picard tools (version 2.27.4; broadinstitute.github.io/picard).

Relative copy number (RCN) profiles were generated using QDNAseq^30^, applying 50 kb binning, GC-content correction, and copy-number segmentation. Segmented bins were annotated with chromosome-arm coordinates using BEDTools and UCSC hg38 cytoband tracks. For each sample, the median log₂ RCN value was computed for each arm (1p-22q), yielding 39 arm-level features derived from ∼49,000 genomic bins. These arm-level summaries were used for downstream statistical analyses and modelling.

### Genome-wide copy number alteration burden

Genome-wide copy number alteration burden (GW-CNA burden) was defined as the proportion of genomic segments with significant copy number deviation.

For Capsule sponge libraries, thresholds for deviation were calculated per chromosome arm using healthy control samples. For each arm, the median ± 1.5× interquartile range (IQR) of log₂-transformed RCN values in controls was computed, and segments outside this range were classified as aberrant. GW-CNA burden for each sample was calculated as:

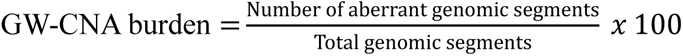

For archival ESD tissue libraries, thresholds were recalibrated using ST-NC samples from the same patient cohort as the diploid reference. The same Tukey outlier approach (median ± 1.5× IQR per arm) was applied.

### Statistical analysis

All statistical analyses were performed in R v4.2.2 (R Foundation for Statistical Computing, Vienna, Austria). Two-sided p ≤ 0.05 was considered statistically significant.

Clinical and demographic variables were summarised using descriptive statistics (mean, median, standard deviation, interquartile range [IQR]).

For Capsule sponge data, GW-CNA burden was compared across endoscopic outcome categories (normal, benign, IEN/early OSCC) using the Kruskal-Wallis test. When significant, pairwise differences were assessed with Dunn’s test and Hochberg correction.

For archival tissue, GW-CNA burden and arm-level RCN values (2q, 3q, 9p, 11q) were compared across histological classes (ST-NC, ANSE, LG-IEN, HG-IEN, OSCC). Global differences were assessed with Kruskal-Wallis followed by Dunn’s test (Hochberg-adjusted). To assess stepwise transitions (e.g. ST-NC→ANSE; ANSE→LG-IEN; LG-IEN→HG-IEN), paired Wilcoxon signed-rank tests were applied. P values were adjusted using the Benjamini-Hochberg method. Alteration frequency for each chromosome arm was defined as the proportion of samples in each class exceeding that arm’s outlier threshold.

### Predictive modelling

Diagnostic models were trained to classify Capsule sponge libraries as positive (IEN/early OSCC) versus negative (normal squamous epithelium or benign/reactive changes not associated with OSCC progression, including oesophagitis, papilloma, acanthosis, necrosis, or koilocytosis). Candidate genomic predictors were derived using elastic net logistic regression (glmnet) on 39 chromosome arm-level RCN features plus GW-CNA burden. The α parameter (L1/L2 ratio) was varied from 0 to 1 in increments of 0.1, and the λ₁se value, corresponding to the largest λ within one standard error of the minimum cross-validation error, was selected to favour parsimonious models. Predictor robustness was evaluated by leave-one-out refitting across all samples. Predictors with non-zero coefficients in ≥70% of iterations and consistent relative risk estimate across refits were retained. This procedure yielded a fixed genomic panel consisting of arm-level relative copy number at 2q, 3q, 9p, and 11q, plus genome-wide CNA burden.

Using these predictors, multivariable logistic regression models (binomial link; caret) were fit under three specifications:

i. **Genomic (continuous):** continuous GW-CNA burden and continuous arm-level RCN (2q, 3q, 9p, 11q);
ii. **Genomic (binary):** binary GW-CNA burden (high vs low, defined a priori) plus the same arms;
iii. **Pathology:** categorical cytology (definite squamous atypia consistent with IEN) and p53 IHC status.

Performance was assessed using stratified repeated cross-validation (5 folds × 10 repeats). Within each resample, Youden’s J statistic defined the operating threshold. Sensitivity, specificity, positive predictive value (PPV), negative predictive value (NPV), F1 score, and accuracy were computed at this threshold. Fold-level metrics were averaged across resamples, and 95% confidence intervals were calculated across folds. Receiver operating characteristic (ROC) curves were generated from pooled out-of-fold predictions, and area under the ROC curve (AUC) with 95% confidence intervals was estimated using the DeLong method (pROC).

## Data Availability

All data produced in the present study are available upon reasonable request to the authors

## Data and code availability

Upon publication, raw sequencing data will be published in access control databases.

Minimal code required for replication of results is provided as supplementary file 1 and 2 and will be released on GitHub upon publication.

## Notes

Conflict of interest: RCF and MoD are named on patents for Cytosponge and related assays licensed to Covidien (now Medtronic). RCF and MoD are co-founders and shareholders of Cyted Ltd. SK is an employee of Cyted Ltd. Cyted had no role in the design or analysis of this study. Remaining authors have no conflict of interest to disclose regarding this report.

### Competing Interest Statement

RCF and MoD are named on patents for Cytosponge and related assays licensed to Covidien (now Medtronic). RCF and MoD are co-founders and shareholders of Cyted Ltd. SK is an employee of Cyted Ltd. Cyted had no role in the design or analysis of this study. Remaining authors have no conflict of interest to disclose regarding this report.

### Funding Statement

The study was funded by the National Science Centre grant OPUS-19 (2020/37/B/NZ5/04003).

